# Real-world effects of ACE inhibitors and Angiotensin Receptor Blockers: protocol for an emulation study of the ONTARGET trial using electronic health records

**DOI:** 10.1101/2021.03.15.21251990

**Authors:** Paris Baptiste, Angel YS Wong, Anna Schultze, Marianne Cunnington, Johannes FE Mann, Catherine Clase, Clémence Leyrat, Laurie Tomlinson, Kevin Wing

## Abstract

**Introduction:** Cardiovascular disease (CVD) is a leading cause of death globally, responsible for nearly 18 million deaths worldwide in 2017. Medications to reduce the risk of cardiovascular events are prescribed based upon evidence from clinical trials which explore treatment effects in an indicated sample of the general population. However, these results may not be fully generalisable because of trial eligibility criteria that generally restrict to younger patients with fewer comorbidities. Therefore, evidence of effectiveness of medications for groups underrepresented in clinical trials such as those over 75 years, from ethnic minority backgrounds or with low kidney function may be limited.

The ONTARGET trial studied the effects of an angiotensin-converting-enzyme (ACE) inhibitor and an angiotensin II receptor blocker (ARB) separately and in combination on cardiovascular event reduction. Using individual anonymised data from this study, in collaboration with the original trial investigators, we aim to investigate clinical trial replicability within routinely-collected patient data. If the original trial results are replicable, we will assess treatment effects and risk in groups underrepresented and excluded from the original clinical trial.

**Methods and analysis:** We will develop a cohort analogous to the ONTARGET trial within CPRD between 1 January 2001 to 31 July 2019 using the trial eligibility criteria and propensity score matching. The primary outcome, as in the trial, is a composite of cardiovascular death, non-fatal MI, non-fatal stroke and hospitalisation for congestive heart-failure, examined in a time-to-event analysis. If results from the cohort study fall within pre-specified limits, we will expand the cohort to include those with advanced kidney dysfunction and increase the proportion of elderly participants and those from ethnicity minority backgrounds.

We will then compare the risk of adverse events and association with long-term outcomes in the clinical trial, with that seen in a directly comparable sample of those attending NHS primary care.

**STRENGTHS AND LIMITATIONS:** *Strengths:*

- Large cohort study giving power to look at effects within subgroups underrepresented in the clinical trial
- Access to individual patient level data from a landmark trial to support creation of a trial-analogous cohort
- Novelty of studying treatment effects of dual therapy in real-world settings

*Limitations:*

- There may be differences between the trial population and the observational cohort due to the level of detail on inclusion/exclusion criteria provided by the trial
- Drug-specific effects are unlikely to be able to be investigated due to small numbers in the dual-therapy arm: class-specific effects will be studied instead
- Misclassification by primary care coding may lead to inaccurate replication of trial inclusion and exclusion criteria.

## INTRODUCTION

Hypertension, age, diabetes and poor diet contribute to cardiovascular disease (CVD), a leading cause of death worldwide [1]. Men have a higher incidence than women, despite women having higher mortality [2]. Angiotensin-converting-enzyme inhibitors (ACE inhibitors) and/or angiotensin II receptor blockers (ARBs) reduce blood pressure by targeting the renin-angiotensin system (RAS). They are commonly used drugs for the treatment of hypertension, stroke, heart failure, other CV outcomes and proteinuric kidney disease [3].

Evidence underpinning the use of ACE inhibitors and ARBs comes from the results of landmark clinical trials. Although these international trials include a large number of participants, many have limited inclusion of subgroups, such as elderly patients, those from ethnic minority groups and people with impaired renal function, and thus have limited power to look for interactions in drug effects [4]. Activity of the RAS and response to drugs that inhibit this system differ between patients, for example among different genders and ethnic groups [5]. In the management of hypertension, there is a longstanding theoretical model that people of black African or African-Caribbean family origin, (subsequently referred to as ‘black’) have lower levels of renin and that some drugs which block the RAS such as ACE inhibitors and ARBs are less effective in black populations [6]. Despite the evidence supporting this, it is increasingly recognised that there are no clear genetic causes of underlying health differences between ethnic groups, and differences may be due to factors such as differences in socio-economic status and access to healthcare, indicating a level of underlying structural racism [7]. Poor representation of black populations in clinical trials limits the ability to examine variation in drug effects by ethnicity [8]. Information regarding drug effects in these underrepresented populations is frequently only available from non-interventional studies, often limited to select patient groups or heavily confounded. Trial-emulation is a technique which can be used to address this issue. By creating a (“trial-analogous”) observational cohort that has similar characteristics to a trial population that has been randomised, and accounting for confounders using propensity score methods, residual confounding can be reduced. If analysis of the trial-analogous cohort generates similar results to the trial, this demonstrates the validity of the patient selection and propensity score methods used. These methods can then be applied to the analysis of the same treatment effects in populations who would have been excluded or underrepresented in the original trial, and populations who are undergoing treatment for a longer period than the follow-up period of the original trial.

We aim to use trial emulation and explore the validity of our methods for assessing treatment effectiveness and risk in non-interventional settings by matching individual patient data from the Ongoing Telmisartan Alone and the Ramipril Global Endpoint Trial (ONTARGET) to a trial-analogous cohort developed in UK primary care data.

We will then apply our validated methods to the estimation of:

1. treatment effects and risk in groups that were excluded from the trial due to age and prior comorbidities, and
2. treatment effects in people over 75 years, of black/Asian ethnicity, those with low kidney function and females who were underrepresented in the trial.

## AIMS AND OBJECTIVES

### Aims

1. To measure the association between ACE inhibitors and ARBs and cardiovascular outcomes within a trial-analogous cohort and within patients excluded and underrepresented in the ONTARGET trial.
2. To help develop a methodological framework that can be used to validate routinely-collected data against randomised controlled trials (RCTs) and obtain conclusions regarding treatment effectiveness and risk caused by treatments in the general population.

### Objectives

1. To explore methods for measuring the effects of ACE inhibitors and ARBs by comparing measures of effect obtained from studies using an RCT-analogous cohort from UK routine primary care with those obtained from a randomised clinical trial;
2. To explore the validity of using electronic health record data to draw conclusions on treatment effectiveness and risk in patients excluded from trials;
3. To explore the validity of using electronic health record data to draw conclusions on treatment effectiveness and risk in patients underrepresented in trials;
4. To investigate long-term outcomes and adverse events of patients treated with ACE inhibitors or ARBs beyond the duration of trials.

## METHODS AND ANALYSIS

### Study design

A cohort design will be used with a trial-emulation component.

### Settings/data sources

Data used in the study will be obtained from the RCT, ONTARGET, and the UK Clinical Practice Research Datalink (CPRD) GOLD (linked to Hospital Episode Statistics database and Office for National Statistics data).

### ONTARGET

The global landmark ONTARGET trial compared the non-inferiority of an ARB (telmisartan 80 mg daily) with an ACE inhibitor (ramipril 10 mg daily), and the superiority of a combination of both therapies compared with ramipril alone [9]. Patients had established vascular disease or were at high risk of vascular disease.

The primary outcome was a composite of: cardiovascular related death, non-fatal myocardial infarction (MI), non-fatal stroke or hospitalisation for heart-failure [10]. Some baseline characteristics are displayed in **Table 1**.

**Table 1.** Baseline characteristics from ONTARGET trial. n (%)= number (percent). Ethnic group was self-reported

| **Characteristic** | **Ramipril**<br>**(N=8576)** | **Telmisartan**<br>**(N=8542)** | **Combination**<br>**Therapy (N=8502)** |
| --- | --- | --- | --- |
| Age – years | 66.4± 7.2 | 66.4 ± 7.1 | 66.5 ± 7.3 |
| Female sex – n (%) | 2331 (27.2) | 2250 (26.3) | 2250 (26.5) |
| Ethnic group – n (%) |  |  |  |
| Asian | 1182 (13.8) | 1172 (13.7) | 1167 (13.7) |
| Arab | 102 (1.2) | 106 (1.2) | 106 (1.2) |
| African | 206 (2.4) | 215 (2.5) | 208 (2.4) |
| European | 6273 (73.1) | 6213 (72.7) | 6222 (73.2) |
| Native or<br>aboriginal | 747 (8.7) | 756 (8.9) | 728 (8.6) |
| Other | 64 (0.7) | 77 (0.9) | 69 (0.8) |
| Missing | 2 (<0.1) | 3 (<0.1) | 2 (<0.1) |

In the intention-to-treat (ITT) analysis, the trial found that telmisartan was non-inferior to ramipril in prevention of the primary composite outcome (RR=1.01, 95% CI: 0.94-1.09) but was less likely to cause angioedema. In addition to this, it showed that combination therapy was no better than ramipril alone (RR=0.99, 95% CI: 0.92-1.07) in preventing the primary composite outcome and significantly increased the risk of hypotension, syncope, renal dysfunction and hyperkalaemia.

Based on the findings of this trial and a smaller parallel trial, TRANSCEND, in October 2009 telmisartan was approved for cardiovascular risk reduction in patients intolerant of ACE inhibitors, aged >55 years and with a high-risk of cardiovascular events, after already having been approved as an antihypertensive drug [11].

### CPRD

CPRD is an anonymised database of patient data from GP practices across the UK. The data consist of 50 million patients with records dating back to 1987, of whom 14 million are currently registered at practices in the UK, ~20% of the UK population [12]. Patients have a median follow-up time of 10 years.

The database contains demographic data, diagnoses and symptoms along with drug exposures, tests and vaccines. Linkage to Hospital Episode Statistics (HES) and other databases such as cancer registries and death registries from the Office for National Statistics (ONS) is also available. In August 2019 linkage data was available from ~74% of CPRD GOLD practices located in England and ~50% of practices in the UK, with 10,800,187 patients eligible for linkage [13].

### Study population

Participants from CPRD with a prescription for an ACE inhibitor or ARB and eligible for HES linkage between 1 January 2001 and 31 July 2019 will be selected. To increase power, we will examine effects of drug classes, rather than specific drugs. Prevalent users were included in the trial, and we will also include patients with previous prescriptions for ACE inhibitors or ARBs. Further detail related to the selection of participants for each objective is provided below.

#### Objective 1

To explore non-interventional methods for measuring the effects of ACE inhibitors and ARBs by comparing results obtained from studies using a non-interventional RCT-analogous cohort from UK routine primary care with those obtained from a randomised clinical trial.

For this objective, users of ARBs will be compared to users of ACE inhibitors.

#### Step 1: Selection of exposed time periods

Prescriptions for an ACE inhibitor or ARB received at least 12 months after the patient has been registered with a general practice that meet prespecified standards for research-quality data (i.e., be ‘up-to-standard’) for at least 12 months will be considered as exposed time periods. Exposed time periods will be defined as periods of continuous therapy i.e., receiving a repeat prescription, >90 days without a prescription after the previous prescription ending will result in the exposure period ending. Prescription duration will be calculated using quantity and daily dose. If this is missing, the median will be imputed. Patients can contribute more than one exposed time period for each drug, with the earliest prescription in each exposed time period denoted as the first eligible prescription.

#### Step 2: Application of inclusion criteria

Exposed time periods where patients are aged ≥55 years and ever received a diagnosis of one of the following prior to the first eligible prescription will be included. This represents the inclusion criteria used in the trial.

- Aged ≥55 years
- At least one of the following of:
  - Coronary artery disease
  - Peripheral artery disease
  - Cerebrovascular disease
  - High-risk diabetes (defined by evidence of end-organ damage)

#### Step 3: Application of exclusion criteria

The trial exclusion criteria will then be applied and time periods with any of the following exclusion criteria prior to the first eligible prescription will be excluded:

- Symptomatic heart failure
- Significant valvular heart disease
- Pericardial constriction
- Complex congenital heart disease
- Uncontrolled hypertension (BP >160/100)
- Elevated potassium above 5.5mmol/L
- Heart transplant recipient
- Stroke due to subarachnoid haemorrhage
- Significant renal disease (defined as patients with codes for renal artery stenosis or renal artery atherosclerosis; or serum creatinine concentration above 265μmol/L)
- Hepatic dysfunction
- Primary hyperaldosteronism
- Hereditary fructose intolerance
- Other major noncardiac illness expected to reduce life expectancy or interfere with participation (cancer, drug or alcohol dependence, mental illness)
- Elevated potassium
- Hypotension

Further information of how these criteria will be interpreted in electronic health records (EHR) is available in the supplementary material and code lists are available for download: https://doi.org/10.17037/DATA.00002112. Due to some of the criteria not being fully assessable using CPRD medical diagnosis codes and laboratory test codes, exclusion criteria are analogous with ONTARGET criteria but we acknowledge they are not identical.

Periods where all inclusion and exclusion criteria are met will be referred to as trial eligible periods and the start date of these periods will be denoted as the eligible for trial inclusion date. The ACE inhibitor exposed cohort will include those periods where a prescription for an ACE inhibitor was received. The ARB exposed cohort will include those periods where a prescription for an ARB is received.

#### Step 4: Matching to trial participants

Having obtained individual patient data for ONTARGET participants, we will match patients within the ONTARGET ramipril study arm to the CPRD ACE inhibitor trial eligible exposure period with the closest propensity score for the probability of being included in the trial. Variables for the propensity score will be chosen based on those known or suspected to influence the likelihood of the outcomes of interest (see Covariates section for further details).

Exact selection of matching variables will depend on the quality and completeness of the data available. Characteristics will be measured at the eligible for trial inclusion date for the ACE inhibitor trial eligible period. Once a trial participant is matched to an ACE inhibitor exposure period from CPRD all other ACE inhibitor exposure periods in CPRD for that participant will be dropped, ensuring a patient can only be matched and included once in the resulting ACE inhibitor trial-analogous cohort. We will consider using methods such as coarsened exact matching and propensity score matching, further details are given in the statistical analysis section. We anticipate matching all or the majority of ONTARGET participants to a CPRD ACE inhibitor-exposed patient, giving us a pool of ONTARGET analogous ACE inhibitor-exposed patients, with similar baseline characteristics to the trial participants at the point of randomisation. This step is outlined in **Figure 1**.

**Figure 1.**
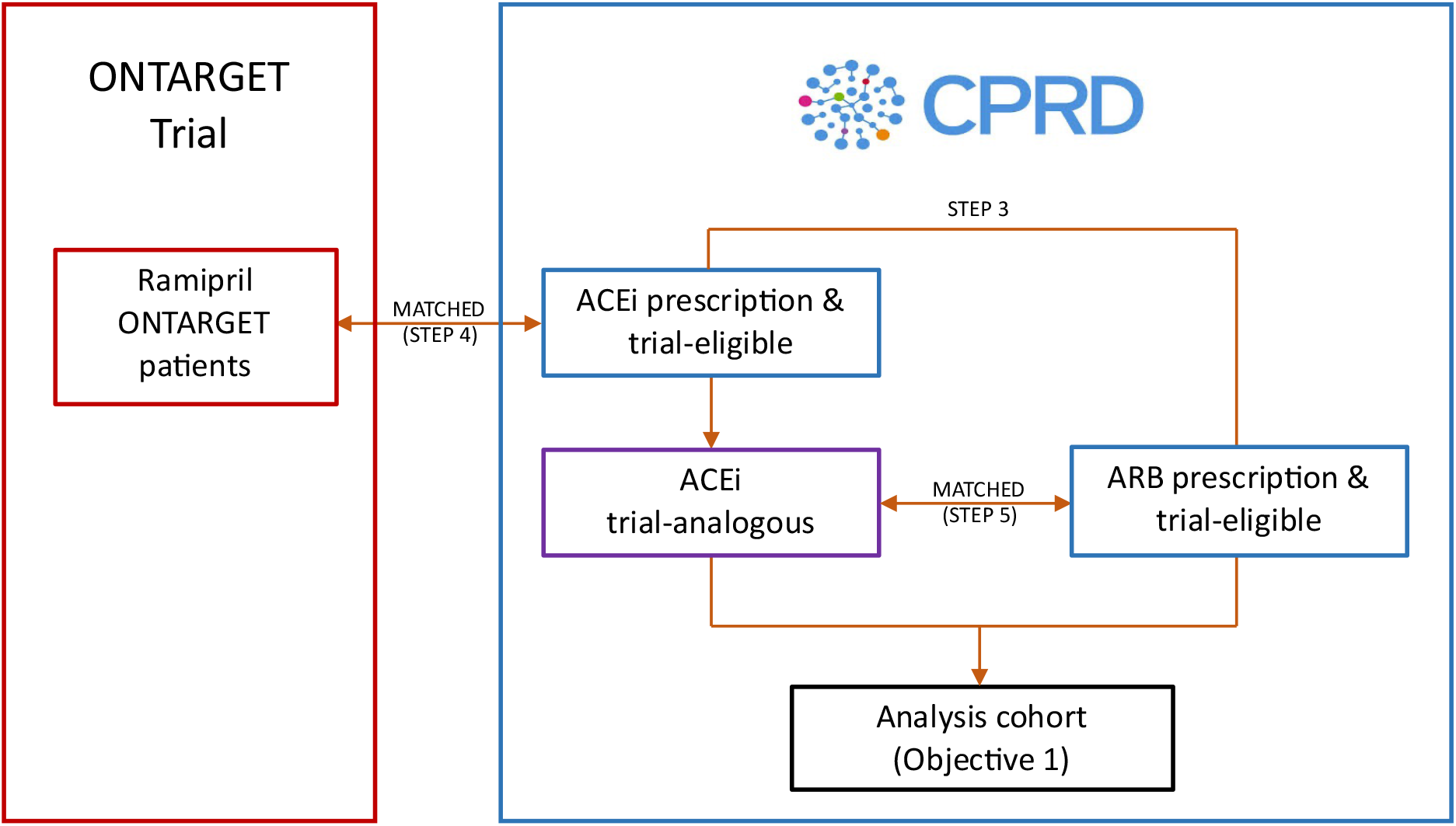
Simplified flow chart illustrating the planned steps in the selection of CPRD patients required to address objective 1. Note double ended arrows denoted ‘MATCHED (STEP X)’ indicates where two cohorts will be 1:1 matched using propensity score matching or some other similar method. CPRD, UK Clinical Practice Research Datalink; ACEi, angiotensin-converting-enzyme inhibitor; ARB, angiotensin II receptor blocker.

#### Step 5: Matching trial-eligible exposure groups

The ACE inhibitor trial-analogous patients selected by Step 4 will be matched 1:1 to the ARB trial-eligible periods from Step 3 with the closest propensity score considering the same variables considered for the propensity score model in Step 4. This matching step will ensure the ARB trial-eligible group has similar characteristics to the telmisartan ONTARGET group due to randomisation in the trial. It will also help us to understand whether trial outcomes can be investigated in non-interventional settings alone, when access to the trial data is not available. Once an ARB exposure period has been matched, any other ARB exposure periods for that patient will be excluded so an ARB patient is matched only once. If a patient ends up contributing eligible exposure periods to both the ARB and ACE inhibitor groups, a restriction will be added that the patient cannot be matched to themselves.

The matched ACE inhibitor and ARB groups from Step 5 will be the analysis cohort for the validation step.

Prior to the remaining objectives, we will check our findings from the validation step are generalisable to other settings. To do this we will repeat this step, matching the ACE inhibitor trial-analogous patients to the dual therapy trial-eligible group and see if results for the primary outcome are comparable with the trial. Dual therapy will be defined as explained in Objective 2.

#### Objective 2

To explore the validity of using electronic health record data to draw conclusions on treatment effectiveness and risk in patients excluded from trials

Those patients who have one of the diagnoses listed in the trial diagnosis criteria in Step 2, but who would have been excluded from the trial due to meeting specific exclusion criteria, such as those: (1) age <55 years or (2) with significant renal disease. Exposure groups will be selected as in Steps 1-3 with the inclusion/exclusion criteria modified to reflect that people either <55 years or with significant renal disease can be included. As the CPRD cohorts will include patients excluded from the trial, the cohorts will not be matched to the trial participants. The propensity score model developed in Step 4 will be the basis for addressing confounding as validated in Objective 1.

Due to the difficulty of defining the dual therapy arm using routine data we will define dual ACE inhibitor/ARB users as patients with overlapping prescriptions who receive an additional prescription for the 1^st^ agent after the 2^nd^ prescription for the 2^nd^ agent, this is shown in **Figure 2**. Follow-up will then be started from the date of the 1^st^ prescription of the 2^nd^ agent, with a sensitivity analysis planned where follow-up starts from the 2^nd^ prescription for the 2^nd^ agent (to evaluate the impact of using a prescription event occurring in the future for defining dual therapy users in the main analysis).

**Figure 2.**
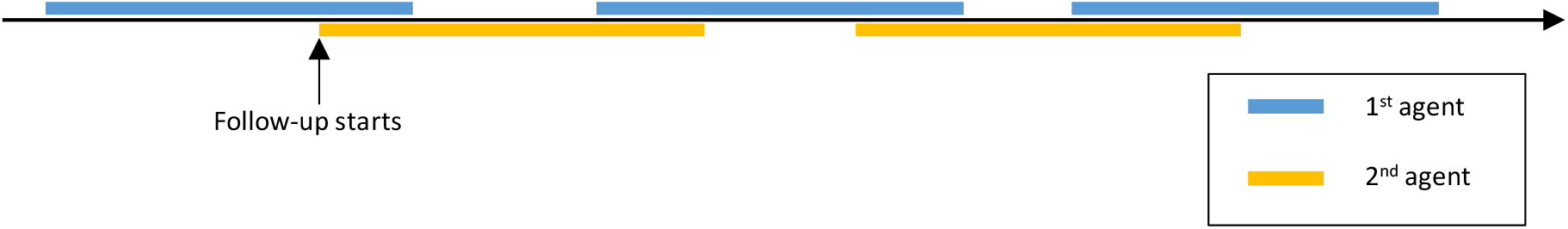
Example timeline of dual therapy user with overlapping prescriptions for 2 agents with follow-up starting at date of 1^st^ prescription for 2^nd^ agent.

#### Objective 3

To explore treatment effectiveness and risk in groups underrepresented in trials.

This will be applied as in Objective 2, with a focus on the groups: (1) of black/Asian ethnicity, (2) aged >75 years, who were underrepresented. All arms will be studied.

#### Objective 4

To investigate long-term outcomes and adverse events of patients treated with RAS blockades beyond the duration of trials.

Adverse events such as cough, angioedema and renal impairment will be studied over a longer duration than that in the trial. This will be studied in the same cohort developed in Step 5 to address Objective 1.

## EXPOSURES, OUTCOMES AND COVARIATES

### Exposures

Exposures will be determined using prescribing records in CPRD and code lists developed for ACE inhibitors and ARBs.

For Objective 1, ARBs are the primary exposure and will be compared to ACE inhibitors. All doses will be considered. If our results show differences to those seen in the trial, we will further explore the drug specific comparison of telmisartan vs. ramipril at all doses for the primary outcome and also when investigating adverse effects.

For the remaining objectives, dual therapy will also be considered as an exposure compared to ACE inhibitors, and will be defined as explained in the ‘study population’ section.

### Outcomes

#### Outcomes to be measured are

- Primary outcome: composite of cardiovascular death, non-fatal MI, non-fatal stroke or hospital admission for congestive heart failure
- Secondary outcomes:
  - Components of primary outcome: (separately) cardiovascular death; non-fatal MI; non-fatal stroke; hospital admission for congestive heart failure
  - (separately) newly diagnosed congestive heart failure; revascularization procedures; nephropathy (defined as 1. 50% reduction in eGFR or start of renal replacement therapy or GFR<15ml/min (for sensitivity analysis requires 50% reduction in eGFR on two occasions at least 3 months apart) and 2. Development of estimated GFR<15 or start of renal replacement therapy (for sensitivity analysis requires eGFR<15 on two occasions at least 3 months apart))
- Other outcomes: (separately) All-cause mortality or microvascular complications of diabetes mellitus.
- Safety outcomes: Cough, angioedema, hyperkalaemia or renal impairment

Outcomes will be identified using read codes and ICD-10 codes in CPRD and HES. Code lists are available for download: https://doi.org/10.17037/DATA.00002112.

### Covariates

The propensity score models in Step 4 and Step 5 of the ‘study population’ section will consider a large range of variables including the following ONTARGET baseline characteristics:

- Age
- Sex
- Ethnicity
- Cardiovascular disease (categorised into-coronary, peripheral, cerebrovascular)
- Diabetes
- Prior treatment with RAS blockers
- Baseline systolic and diastolic blood pressure within 6 months
- Smoking status
- Body mass index (BMI)
- Renal function

In the propensity score model in Step 5 of the ‘study population’ section variables such as calendar period and healthcare utilisation (e.g., GP consultations, hospital appointments, procedures) will also be considered.

### SAMPLE SIZE

We will include all eligible patients registered in CPRD who meet the trial inclusion/exclusion criteria where applicable. In ONTARGET there were 8576 in the ramipril arm, 8542 in the telmisartan arm and 8502 in the combination arm so we estimate a minimum of 14,000 CPRD patients exposed to an ACE inhibitor or an ARB are required for the individual patient matching to provide any benefit.

In a previous study [14] the following counts were obtained: ACE inhibitor alone: n=281,204, ARB alone: n=83,850, both ACE inhibitor and ARB at the same time: n=39,548 between April 1997 and March 2014. Using data from an ongoing study (ISAC Protocol 19_072, using CPRD GOLD alone) we estimate that 37% of ACE inhibitor/ARB users are aged ≥55 years with previous cardiovascular or cerebrovascular disease and/or diabetes at drug initiation.

We have assumed a sample size of 80,000, 20,000 and 14,000 in the ACE inhibitor, ARB and dual therapy groups, respectively. We have chosen sample sizes smaller than those obtained from 37% of the cohort sizes described in the study by Mansfield et Al. [14] since these are more likely to reflect the numbers found after applying the trial exclusion criteria. We have taken the upper and lower confidence limits for the risk ratio for the primary outcome in ONTARGET and the baseline risk of 16.5% in the ramipril group [10]. From this we estimate 87.4% power for a risk ratio of 0.94, and 99.6% power for a risk ratio of 1.09, when comparing the non-inferiority of ARBs vs. ACE inhibitors. For the superiority of dual therapy vs. ACE inhibitors, we estimate 94.6% power for a risk ratio of 0.92, and 87.0% power for a risk ratio of 1.07.

## STATISTICAL ANALYSIS

### Propensity score for addressing confounding

Multivariable logistic regression (on probability of being included in the trial for Step 4, and on exposure status for Step 5) will be used to generate the propensity score, with the variables selected for inclusion in the initial multivariable logistic regression model based upon expert/prior knowledge of association with outcome. Those provisional variables listed in the ‘Covariates’ section along with other variables will be considered.

The propensity score model developed in the validation step in Objective 1 will be the basis for the model used in Objectives 2 and 3.

## Methods of analysis

For Objective 1 comparisons between ACE inhibitors and ARBs will be made, and a comparison among ACE inhibitors and dual therapy for the primary outcome only. For the other objectives the comparison between ACE inhibitors and ARBs will be made in addition to the comparison of ACE inhibitors alone and dual therapy. A sensitivity analysis will be carried out to compare drug-specific effects of ramipril and telmisartan, if results differ from the trial. An intention-to-treat (ITT) analysis will be carried out for the validation of results in Objective 1, which was used in ONTARGET [10] and the remaining objectives.

For Objectives 2-4 a per-protocol (PP) analysis will be carried out (in addition to ITT) for all comparisons. Patients who discontinue or switch treatment or start dual therapy, data for original treatment will be included up to and including their calculated date of last dose of the initially prescribed treatment + 60 days, to account for repeat prescriptions and ensure exposure groups are correctly categorised. Their exposure period for the new treatment will start from the date of first prescription for new drug. Therefore, patients may contribute more than one exposure period. The two analysis populations are shown in **Figure 3**. Patients will be censored up to the earliest of: outcome of interest, death, leaving general practice date, or last data collection date from the general practice, or the derived date of last dose of study drug when using the PP analysis. If these dates do not occur the patient will be censored after 5.5 years of follow-up (reflecting the maximum follow-up time in the trial). Due to patterns of use of these drugs in EHR, we anticipate dual therapy users will have initially been single therapy users introducing some bias. We hope to address this by comparing the ITT and PP results.

**Figure 3.**
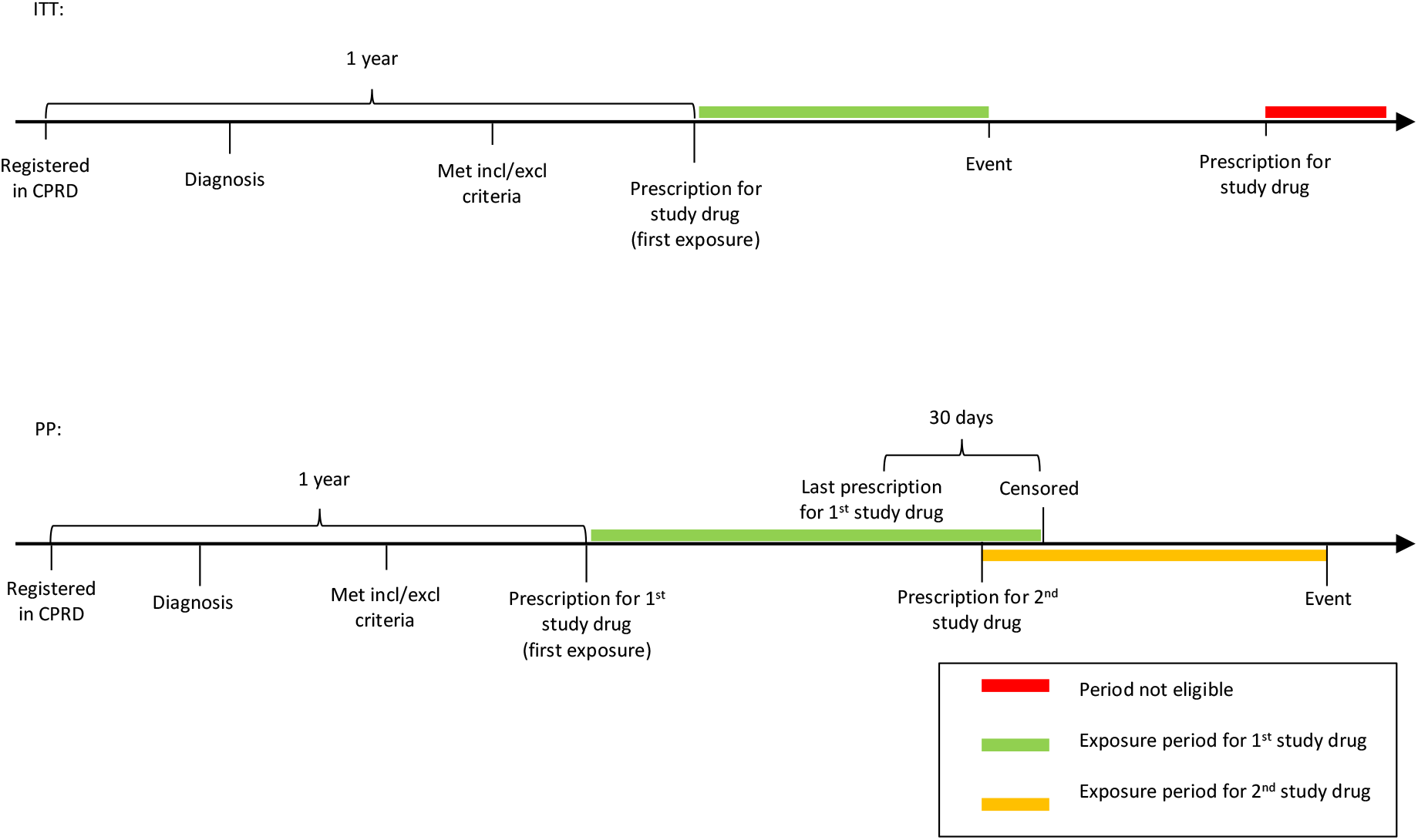
Figure illustration analysis groups to be used to address objectives. ITT timeline demonstrates order that criteria must be met for exposure period to be eligible, with patient no longer being able to contribute additional expose periods after being censored. PP timeline shows in green where patients exposure period can contribute to exposure group 1, then in yellow where a patient switches treatment and can contribute to second exposure group. There will be a small period of overlap, where the patient will contribute to both exposure groups as shown in the figure. ITT, intention-to-treat; PP, per-protocol.

Our analysis will mirror the approaches used in ONTARGET. A Cox proportional hazards model will be used to address the primary composite outcome of time to cardiovascular death, non-fatal myocardial infarction, non-fatal stroke, or hospitalisation for congestive heart failure. Point estimates and two-sided 95% confidence intervals for hazards ratios will be provided for all efficacy outcomes with the bootstrap method used to estimate standard errors. Other outcomes and safety outcomes will be studied using logistic regression. If variability between practices is observed, a mixed effects model will be considered to account for this. A summary table of our protocol compared to the ONTARGET protocol is given in **Table 2**.

**Table 2.** Table of key design aspects of the ONTARGET trial and how these will be interpreted in our CPRD cohort.

| Protocol component | Description in ONTARGET | Description in CPRD |
| --- | --- | --- |
| Eligibility criteria | Patients aged $\geq 55$ years with coronary artery, peripheral vascular, or cerebrovascular disease or high-risk diabetes with end organ damage recruited up to 2004. No restriction on previous ACE inhibitor/ARB use except must be able to discontinue use. | Patients with a prescription for an ACE inhibitor or ARB between 01 January 2001 to 31 July 2019, eligible for HES linkage, aged $\geq 55$ years with coronary artery, peripheral vascular, or cerebrovascular disease or high-risk diabetes. |
| Treatment strategies | Patients will enter 3-week single blind run-in period to check compliance then will be randomised to one of the 3 trial arms: ramipril 10mg + telmisartan placebo, telmisartan 80mg + ramipril placebo or ramipril 10mg + telmisartan 80mg. | Continuous courses of therapy with treatment gaps of $< 90$ days. Dual therapy users defined as patients with overlapping prescriptions who receive additional prescription for the 1<sup>st</sup> agent after the 2<sup>nd</sup> prescription for the 2<sup>nd</sup> agent. |
| Assignment procedures | Randomly assigned and will receive a placebo for other drug so unaware which arm they are assigned to. | Based on prescriptions received.<br>Patient can contribute to all three exposure groups at different timepoints. |
| Follow-up period | Follow-up starts at randomisation and ends at primary event, death, loss to follow-up or end of study.<br>Close out planned in July 2007 | Follow-up starts at start of trial-eligible period where exposure period meets trial inclusion/exclusion criteria. Ends at the earliest of: outcome of interest, death, transferred out of practice date, or last data collection from the general practice. If these dates do not occur the patient will be censored after 5.5 years of follow-up. |
| Outcome | Primary composite outcome of: cardiovascular death, non-fatal MI, non-fatal stroke, hospitalisation for heart failure | Primary composite outcome of: cardiovascular death, non-fatal MI, non-fatal stroke, hospitalisation for heart failure |
| Analysis plan | Primary analysis time-to-event counting first occurrence of any component of the composite outcome using Cox proportional hazards model. | Match to trial to obtain trial-analogous cohort then will match trial-eligible exposure groups.<br>Cox proportional hazards model will be used for primary analysis. |

### Validation of results against ONTARGET

In Objective 1 alone we will validate the findings from our primary analysis against ONTARGET by determining whether results of the CPRD analysis are comparable with the ONTARGET trial results. The ONTARGET trial demonstrated non-inferiority of telmisartan over ramipril for the primary outcome (RR=1.01, 95% CI=0.94-1.09) under an ITT analysis and showed similar results under a PP analysis giving RR=1.00 (95% CI=0.92-1.09) [10].

Since the primary outcome comparing telmisartan vs. ramipril showed clear non-inferiority of telmisartan and the upper limit of the 95% CI was within the non-inferiority boundary of 1.13, this will be used to validate results when testing ARB vs. ACE inhibitors in the CPRD population. To conclude that our results are compatible with the ONTARGET trial results we have two criteria that must be met.

1. Firstly, the effect size for the two exposure groups must be clinically comparable with the ONTARGET findings; the RR for the composite primary outcome (time to cardiovascular death, non-fatal MI, non-fatal stroke, or hospitalisation for congestive heart failure) in the CPRD population under an ITT analysis must be between 0.9 and 1.12.
2. Secondly, the 95% CI for the RR must contain 1.

### Handling measurement of adherence to medication

A sensitivity analysis will be carried out to investigate the effect of a run-in period for compliance. The 3-week run-in period in the trial will be replicated by a 28-day period, reflecting a general prescription duration. Follow-up will be started from 28 days after first prescription and those patients who receive no subsequent prescriptions after 28 days will be excluded.

When using efficacy outcomes for validity we expect different adherence in routine clinical practice compared with the trial. Adherence will therefore be estimated in the CPRD cohort to enable comparisons with the trial and investigate the extent to which this may have influenced any observed differences in treatment effect. We will estimate the proportion of time covered by prescribing as a proxy measure for adherence in CPRD; this proxy measure assumes that all prescriptions are filled and that a patient takes all tablets in the prescription so is although not completely accurate, provides an indication of adherence [15].

### Missing data

CPRD data has few missing data for drug prescribing and mortality (partly through ONS linkage). Information on important comorbidity is also well recorded. Our approach for handling missing data in terms of the baseline characteristics will depend on the variable itself. In some cases, such as for renal function, a patient is more likely to have no data entered if there is no clear clinical evidence of abnormality; in such cases we may instead categorise the variable into a binary parameter e.g., ‘renal dysfunction’ vs ‘no renal dysfunction’. With missing data contributing to the ‘no renal dysfunction’ group. For variables such as BMI and SBP it cannot be assumed that the data are missing at random as we expect that a patient is less likely to have blood pressure recorded if they do not have hypertension. Patients with variables missing that cannot be assumed to be missing at random will therefore be excluded from the trial-eligible cohort prior to Step 4. In cases where missing data can be assumed to be missing at random or missing completely at random both a complete-case analysis and an analysis using multiple imputation in propensity score modelling to impute missing values will be used [16].

## Data Availability

Data was provided by CPRD under terms of the LSHTM license and can be requested from CPRD. ONTARGET trial data was obtained via a data sharing agreement with the sponsor, Hamilton Health Sciences Corporation.

https://www.cprd.com/

## ETHICS AND DISSEMINATION

### Ethics

An application for scientific approval related to use of the CPRD data has been approved by the Independent Scientific Advisory Committee of the Medicines and Healthcare Products Regulatory Agency (protocol no. 20_012). CPRD are already approved via a National Research Ethnics Committee for purely non-interventional research of this type. Access to the individual patient data from the ONTARGET trial was obtained by the trial investigators.

### Dissemination

The results of the study will be submitted to peer reviewed journals and we anticipate three publications to arise directly from the planned work. Findings will also be presented at conferences such as the International Society for Pharmacoepidemiology Conference. Results will also be published on the London School of Hygiene & Tropical Medicine website and in the PhD thesis of the principal investigator. Results that may impact on treatment guidelines will be shared with policy makers such as the Medicines and Healthcare products Regulatory Agency and the National Institute for Health and Care Excellence.

## AUTHORS’ CONTRIBUTIONS

PB, LT and KW contributed to the study question and design. PB wrote the first draft of this protocol based on original scientific approval applications to ISAC that PB, LT, KW, JM, and CC contributed to. PB, LT, KW, AW, CL, MC, AS, JM and CC contributed to further drafts and approved the final version.

