## Supplementary material for "Real-world effects of ACE inhibitors and Angiotensin Receptor Blockers: protocol for an emulation study of the ONTARGET trial using electronic health records"

Table of trial diagnoses (inclusion criteria) and interpretation in CPRD.

| **ONTARGET/TRANSCEND** | **CPRD GOLD (HES + ONS Linked)**  **READ or ICD 10 code for:** |
| --- | --- |
| Aged ≥55 years | Aged ≥55 years prior to prescription of drug |
| Coronary artery disease |  |
| Previous myocardial infarction (>2 days post uncomplicated MI) | MI at least 2 days prior to prescription of drug |
| Stable angina or unstable angina >30 days before informed consent and with documented evidence of multivessel coronary artery disease | Angina/stable angina/unstable angina at least 30 days before prescription of drug and previous coronary artery disease diagnosis |
| Multi-vessel PTCA >30 days before informed consent | Read, ICD-10 or OPCS code for coronary angioplasty at least 30 days before prescription of drug |
| Multi-vessel CABG surgery >4 years before informed consent, or with recurrent angina following surgery | Read, ICD-10 or OPCS code for CABG at least 4 years before prescription of drug or with angina after CABG |
| Peripheral artery disease |  |
| Previous limb bypass surgery or angioplasty | Read, ICD-10 or OPCS code for limb bypass surgery or angioplasty |
| Previous limb or foot amputation | Read, ICD-10 or OPCS code for limb/foot amputation |
| Intermittent claudication, with ankle:arm BP ratio <=0.80 on at least 1 side | Intermittent claudication |
| Significant peripheral artery stenosis (>50%) documented by angiography or non-invasive test | Not applicable |
| Cerebrovascular disease |  |
| Previous stroke | Stroke before prescription of drug |
| Transient ischemic attacks >7 days and <1 year before informed consent | Transient ischemic attacks before prescription of drug |
| High-risk diabetes with evidence of end-organ damage |  |
| High-risk diabetes | Specific codes for diabetes with retinopathy, neuropathy, chronic kidney disease or proteinuria before prescription of drug or diabetes defined by diabetes codes or diabetes therapy with CKD defined as eGFR<60 or proteinuria defined as ACR>3 |

Notes: Where dates are used as criteria dates from both CPRD and HES will be used, but if available HES will be preferred.

| Table of trial exclusion criteria and interpretation in CPRD. | |
| --- | --- |
| **ONTARGET/TRANSCEND exclusion criteria** | **CPRD GOLD (HES + ONS Linked)**  **READ or ICD 10 code (prior to eligible for inclusion date, unless otherwise specified) for:** |
| Inability to discontinue ACE inhibitors or ARB | Not applicable |
| Known hypersensitivity or intolerance to ACE inhibitors or ARB | Not applicable |
| Symptomatic congestive heart failure | Heart failure or left ventricular dysfunction |
| Hemodynamically significant primary valvular or outflow tract obstruction | Aortic or pulmonary stenosis or previous valve replacement |
| Constrictive pericarditis | Constrictive pericarditis |
| Complex congenital heart disease | Congenital heart disease |
| Syncopal episodes of unknown etiology <3 months before informed consent | Not applicable |
| Planned cardiac surgery or PTCA <3 months of informed consent | Not applicable |
| Uncontrolled hypertension on treatment (e.g. BP >160/100 mm Hg) | Last recorded BP >160/100 mmHg for patients on treatment with other antihypertensives prior to ACEI/ARB initiation |
| Heart transplant recipient | Read, ICD-10 or OPCS code for heart transplant recipient |
| Stroke due to subarachnoid haemorrhage | Previous cerebral haemorrhage |
| Significant renal artery disease | Codes for renal artery stenosis or renal artery atherosclerosis; or serum creatinine concentration above 265μmol/L |
| Hepatic dysfunction | Cirrhosis or other documented liver disease |
| Uncorrected volume or sodium depletion | Not applicable |
| Primary hyperaldosteronism | Primary hyperaldosteronism/ Conn’s syndrome |
| Hereditary fructose intolerance | Hereditary fructose intolerance |
| Other major noncardiac illness expected to reduce life expectancy or interfere with study participation | Recorded solid organ or metastatic malignancy within the last 5 years, drug, alcohol dependence or mental illness. |
| Simultaneously taking another experimental drug | Not applicable |
| Significant disability precluding regular follow-up visits | Not applicable |
| Unable or unwilling to provide written informed consent | Not applicable |
| Elevated potassium above 5.5mmol/L | Elevated potassium above 5.5mmol/L |
| Hypotension | SBP <90 mm Hg |
| Notes: Where dates are used as criteria dates from both CPRD and HES will be used, but if available HES will be preferred. Not applicable used when anticipated there will be extensive missing data or risk of misclassification. | |
